# Systematic evaluation of the two main blood-based RNA-seq approaches for Mendelian disease diagnosis

**DOI:** 10.1101/2023.04.22.23288779

**Authors:** Ziying Yang, Xiaoru Yang, Yunmei Chen, Zhonghua Wang, Lijie Song, Jun Sun, Huanhuan Peng, Xunzhe Yang, Zhiyu Peng, Yi Dai

**Affiliations:** College of Life Sciences, University of Chinese Academy of Sciences, Beijing 100049, China; Tianjin Medical Laboratory, BGI-Tianjin, BGI-Shenzhen, Tianjin 300308, China; BGI Genomics, BGI-Shenzhen, Shenzhen 518083, China; Department of Neurology, Peking Union Medical College Hospital, Chinese Academy of Medical Sciences and Peking Union Medical College, Beijing 100730, China

**Author notes:** **Correspondence:** Yi Dai. These authors contributed equally to this work and share first authorship (Ziying Yang, Xiaoru Yang). (Z.Y.). (X.Y.); (Y.C.); (Z.W.); (J.S.). (Z.P.); (H.P.). (Y.D.) (XZ.Y.).

**Keywords:** blood-based RNA-seq, genetic diagnosis, Mendelian diseases, polyA-selection, rRNA depletion

## Abstract

**Background:** As an adjunct to diagnostic exome sequencing and whole-genome sequencing, RNA sequencing (RNA-seq) has been demonstrated to improve diagnostic yield for Mendelian diseases. However, systematic evaluation of the associated experimental and computational processes and the establishment of robust and efficient practices for RNA diagnostics implemented in the clinic to analyse readily accessible whole blood samples are still required.

**Methods:** We simulated clinical conditions in which each patient’s sample is tested only once, and we evaluated the two typical experimental protocols (polyA-selection and rRNA depletion) by comparing the expression profiles, aberrant splicing events and monoallelic expression (MAE) identified from 11 patients in clinical settings with different bioinformatics software.

**Results:** We demonstrated that a higher proportion of unique reads from polyA-selection than rRNA depletion were mapped to exons or exon – intron junction regions (84.54% vs. 40.14%), resulting in more detectable OMIM genes (TPM > 1) in the blood (65.29% vs. 59.79%); thus, the rRNA depletion method requires a median of 258 more valid reads per gene to achieve the same level of gene quantification. Moreover, although the transcriptome profiling of protein-coding genes in the two methods is highly correlated, polyA-selection offers more sensitive detection of MAE variants and aberrant splicing under common filtering conditions in combination with DROP.

**Conclusions:** A combination of polyA+ and DROP is recommended when implementing blood-based RNA-seq for the diagnosis of Mendelian diseases in clinical practice, and filtering criteria for aberrant expression, aberrant splicing and MAE variants are suggested for reference.

## Introduction

Massively parallel technologies provide powerful approaches that, when integrated with sequencing and phenotypic data, contribute compelling information for Mendelian disease diagnosis. The clinical implementation of whole-exome sequencing (WES) and whole-genome sequencing (WGS) has revolutionized genetic diagnostics for individuals suspected to have Mendelian disease by improving diagnostic yield and accelerating the discovery of novel disease genes (1–4). The diagnostic yield of WES ranges from 10% to 70% depending on different cohort scales, phenotypic subgroups, recruitment criteria or test types (5–8), while the diagnostic rate of WGS is 5% greater than that of WES (9); nevertheless, both of these approaches leave a large fraction of patients without a conclusive genetic diagnosis. There are several factors that account for these inconclusive cases. The main challenges are interpreting variants of unknown significance (VUSs) in noncoding regions and identifying all potential disease-causing variants in the coding regions (10, 11). Although the *in silico* prediction tools currently in use perform quite well, additional functional studies involving omics techniques beyond DNA are still needed to verify the biological impacts of specific variants on genes.

RNA-seq can be used to measure the expression levels of thousands of genes simultaneously and identify genes expressed at aberrant levels; it can also provide novel insights into alternative splicing and monoallelically expressed (MAE) rare variants. Recently, to improve the diagnostic yield for genetic disorders, some groups performed blood transcriptome sequencing on patients with negative WES or WGS results and diagnosed an additional 7.5% (12–14), lower than the 10–36% (11, 15, 16) of performing RNA sequencing (RNA-seq) using focal tissues such as muscle and fibroblasts. However, evidence suggests that 70.6% of disease-causing genes in the Online Mendelian Inheritance in Man (OMIM) database are expressed in blood (12), and whole blood is more readily accessible than other tissues, making blood transcriptomics more practical for clinical application. Two main approaches, polyA-selection (polyA+) and rRNA depletion (ribo-minus), are commonly used for protein-coding RNA selection, and the former method has been used in most eukaryotic mRNA studies. In fact, several studies have suggested that certain functional noncoding and protein-coding RNAs do not possess polyA tails, for example, replication-dependent histones (17) and various long noncoding RNAs (lncRNAs) (18, 19). Conversely, ribo-minus libraries captured both polyA+ and polyA-RNAs and a larger proportion of intronic sequences from premature mRNAs and provided a more thorough profile of the transcriptome. However, when using blood transcriptome data to provide evidence for the functional impact of the candidate variants in the context of clinical practice, choosing which method to use to select RNA can have critical effects on subsequent data analysis. The selection method directly determines whether high and uniform coverage of reads in clinically significant OMIM genes is obtained, which is critical for detecting differentially expressed genes and for discriminating variant-associated splicing outcomes between multiple affected individuals in a reproducible and sensitive manner. Although comparisons of the ribo-minus and polyA+ methods have been carried out with different kits and samples since 2010 (20–23), most studies have relied on animal samples or cell-line-derived RNAs, and per-sample biological replicates were performed in almost all studies. In addition, the results tend to reflect the expression profiles of all genes, which is comprehensive and accurate but may lack specific application scenarios. When using the blood transcriptome for the diagnosis of monogenetic disorders in clinical settings, it is critical to focus on alternative splicing events and MAE variants for clinically significant OMIM genes in addition to expression levels. More importantly, for clinical purposes, a patient’s sample can rarely be replicated in one test.

Based on previous studies, we hypothesized that the polyA+ method would be more applicable to Mendelian disease diagnosis in clinical practice. In this paper, we improved upon the commonly used polyA+ method by moving the depletion of globin mRNA from before the oligo(dT) selection step and making it concurrent with the fragmentation step during library generation. To verify our hypothesis and establish standardized practices for RNA diagnostics using clinically accessible whole blood samples, we evaluated the two protocols (ribo-minus and modified polyA+) by comparing the expression profiles, alternative splicing events of OMIM genes and MAE variants identified in clinical settings where RNA samples were collected primarily from patients, most of whom had genome data available, and tested these samples without replication.

## Materials and method

### Patient recruitment and sample collection

This study was approved by the Ethics Committee of Peking Union Medical College Hospital. Eleven patients (7 males and 4 females) with suspected hereditary neuromuscular disorders were recruited, and their whole blood samples were collected in PAXgene Blood RNA tubes. Written informed consent was obtained from all patients.

### RNA extraction

Total RNA was isolated using the PAXgene Blood RNA Kit (Qiagen, Chatsworth, CA, USA) following the manufacturer’s instructions. The yield and quality of the isolated RNAs were assessed using a Qubit fluorometer (Thermo Fisher Scientific, Waltham, MA, USA) and Qsep 100 Bioanalyzer (Bioptic, Taiwan, China), respectively. RNAs with RNA integrity number (RIN) values ≥ 7.0 proceeded to the RNA selection step.

### Ribo-minus library construction

A total of 500 ng of each RNA sample was processed with a Ribo-off Globin & rRNA Depletion Kit (Vazyme, Nanjing, China) for both globin mRNA and rRNA depletion. Briefly, total RNA was hybridized with a mix of globin mRNA oligos and rRNA oligos, and RNase H digestion was performed to deplete rRNA and globin mRNA. To remove the excess single-stranded oligos, DNaseI treatment was carried out after RNase H digestion. Stranded RNA libraries were prepared using a VAHTS Universal V8 RNA-seq Library Prep Kit (Vazyme, Nanjing, China) through fragmentation, reverse transcription, end repair, A-tailing, adaptor ligation and library enrichment.

### PolyA+ library construction

A total of 500 ng of each RNA sample was processed with oligo(dT) beads (Vazyme, Nanjing, China) to select poly A+ RNA. Globin mRNA was removed using a QIAseq FastSelect-Globin Kit (Qiagen, Chatsworth, CA, USA) during the RNA fragmentation step during library preparation. Briefly, mRNA was combined with magnetic oligo(dT) beads, and poly A+ RNA was obtained after separation and purification. Globin mRNA probes were added into the fragmentation system, the probe was hybridized with the globin mRNA sequence through gradient cooling, and reverse transcription was suppressed to remove globin mRNA. Reverse transcription and the following library preparation were consistent with those in the ribo-minus library construction.

### Sequencing and data analysis

Fourteen libraries from each protocol, including 11 patients and 3 replicates of Universal Human Reference RNA (UHRR), were sequenced with a 100 bp paired-end read protocol on MGISEQ-2000 (MGI, Wuhan, China). Approximately 70 M reads (14 Gb raw data) per sample were generated. Sequence data quality was evaluated using FastQC (v0.11.9) combined with MultiQC (v1.11). The STAR (v2.7.9) aligner was used to align clean reads to the human genome (hg19/GRCh37). After alignment, the final BAM files were quantified using HTseq (v1.99.2) by Ensembl annotations (Gencode, GRCh37, release 19) and then normalized to TPM (transcripts per million mapped reads) values using RSEM (v1.3.1). For sample correlation analysis, normalized expression values (ln (TPM + 1)) were used. EdgeR (v3.36.0) and DROP (v1.2.1) were used to detect differentially expressed genes (DEGs), and alternative splicing (AS) events were identified by using Leafcutter (v0.2.9) and rMATS (v4.1.2) with default parameters. Another 10 samples obtained using the same experimental method were used as controls for DEG and AS.

### Reverse transcription polymerase chain reaction (RT–PCR) analysis

The PrimeScript™ II 1st Strand cDNA Synthesis Kit (Takara, Kusatsu, Japan) was used to generate complementary DNA (cDNA) from 500 ng of RNA according to the kit instructions. PrimeSTAR® HS DNA Polymerase (TaKaRa, Kusatsu, Japan) was used for the PCRs. Primers were designed using Primer3Plus (v3.2.6) according to the mRNA sequence and synthesized by Sangon Biotech (Shanghai, China). Control cDNA was obtained from other cases who did not carry the same variants. All PCR products were analysed on a 2% agarose gel. In our study, one or two samples from individuals of the same sex and similar age were used as controls for each group.

## Result

### 1. Structure design and RNA-seq quality control metrics

To fairly evaluate the performance and the impact on clinical interpretation of the RNA-seq data between the two protocols, we simulated the clinical condition where each sample from 11 patients was tested only once for each protocol and 3 replicates of UHRR were used as biological replicates. We used the same bioinformatic pipeline to analyse the sample data one by one by using the remaining sample data from the corresponding protocol as a control. For Mendelian disease diagnosis, the analysis focuses on aberrant expression, aberrant splicing and MAE variants. The study design is depicted in **Figure 1**.

**Figure 1.**
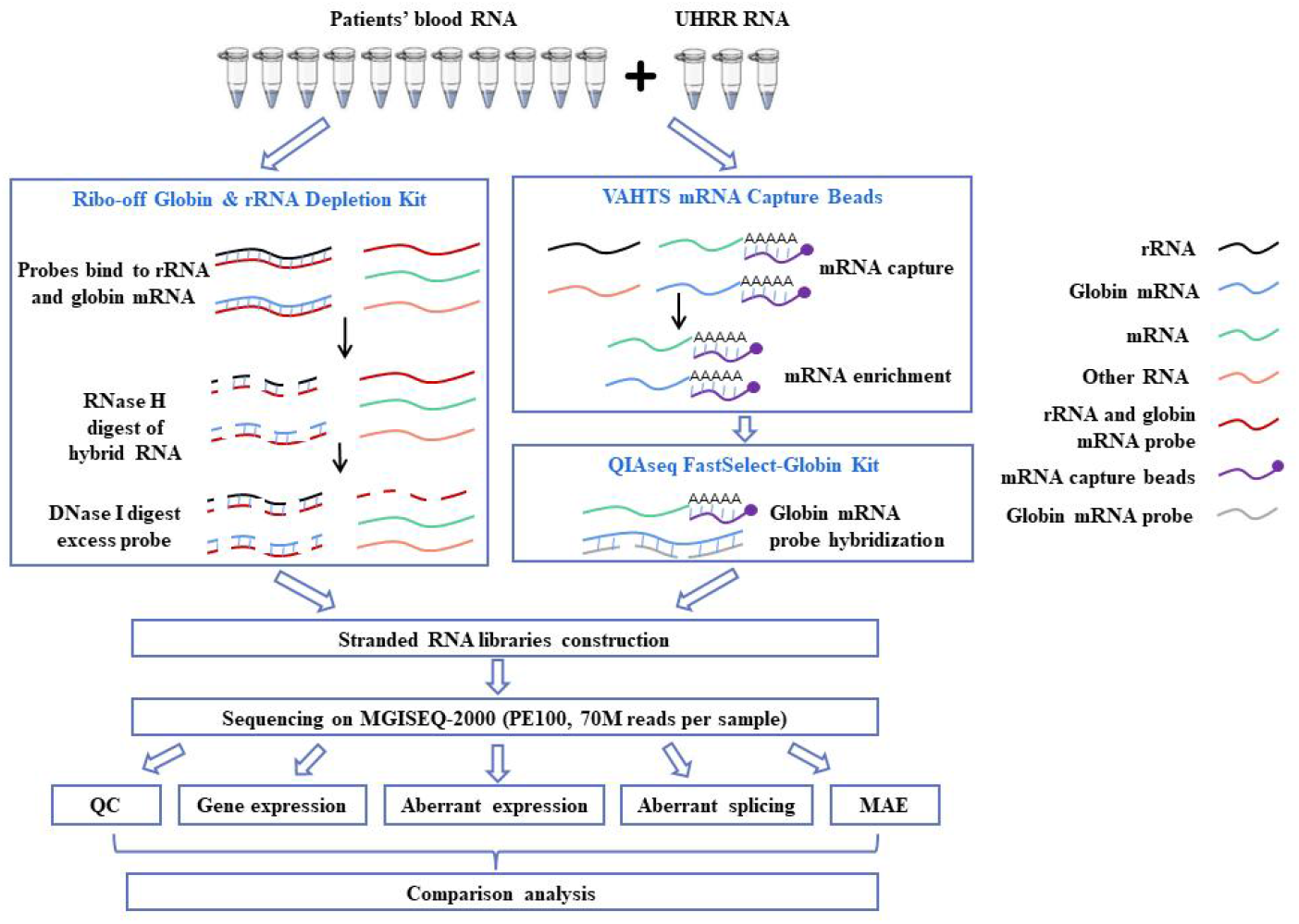
Study design. Blood RNA from 11 patients with suspected hereditary neuromuscular disorders and 3 replicates of Universal Human Reference RNA (UHRR) were performed in parallel by polyA-selection (polyA+) and rRNA depletion (ribo-minus). The expression profiles, aberrant splicing and monoallelically expressed (MAE) rare variants derived from the two methods were compared to determine which method is most suitable for genetic diagnosis and to establish standardized practices for RNA diagnostics using clinically accessible whole blood samples.

In the clinical setting, it is worth paying attention to the available reads covering the target region. The number of total reads and the unique read distribution across different regions from the polyA+ and ribo-minus protocols are listed in **Table 1**. From the polyA+ RNA-seq data, we obtained 19.37 G of raw reads on average per sample, and 94.05% of the high-quality reads were uniquely aligned. We further aligned the unique reads onto RefSeq-defined gene models and found that 66.30%, 10.84%, 18.24%, and 4.61% of them were within exonic, intronic, exon–intron junction, and intergenic regions, respectively. In contrast, from the ribo-minus RNA-seq data, we obtained 20.62 G of raw reads on average per sample, and 92.05% of the high-quality reads were uniquely aligned. We further aligned the unique reads onto RefSeq-defined gene models and found that 29.56%, 51.21%, 10.58%, and 8.65% of them were within exonic, intronic, exon–intron junction, and intergenic regions, respectively. These observations are consistent with those of previous studies showing that reads from the polyA+ library mapped more frequently to exonic and exon–intron junction regions than those from the ribo-minus library. We also compared the globin mRNA and rRNA removal results of the two methods because they may affect the quality and accuracy of the gene expression profiles. Although the read ratio of globin mRNA and rRNA was extremely low in both methods, the ratio in the polyA+ RNA-seq data (0.21% and 2.46%, respectively) was lower than that in the ribo-minus RNA-seq data (0.29% and 3.44%, respectively), which indicated that the removal was superior in the polyA+ protocol.

**Table 1.** Summary of read mapping

| Protocol | Sample | Raw data | Unique<br>mapping<br>Ratio | Exonic<br>reads<br>ratio | Intronic<br>reads<br>ratio | Exon-intr<br>on<br>junction<br>reads ratio | Interg<br>enic<br>reads<br>ratio | rRNA<br>reads<br>ratio | Globin<br>mRNA<br>reads<br>ratio |
| --- | --- | --- | --- | --- | --- | --- | --- | --- | --- |
| PolyA+ | P1 | 18.75 Gb | 95.67% | 63.29% | 15.31% | 17.37% | 4.03% | 0.64% | 0.20% |
|  | P2 | 17.74 Gb | 93.25% | 69.59% | 7.96% | 18.19% | 4.26% | 2.65% | 0.26% |
|  | P3 | 21.48 Gb | 94.03% | 68.32% | 9.83% | 17.63% | 4.22% | 2.20% | 0.26% |
|  | P4 | 12.35 Gb | 95.08% | 67.60% | 11.34% | 17.17% | 3.88% | 1.39% | 0.23% |
|  | P5 | 20.90 Gb | 92.61% | 67.39% | 9.40% | 18.16% | 5.05% | 2.94% | 0.19% |
|  | P6 | 18.98 Gb | 95.01% | 63.33% | 15.28% | 17.00% | 4.39% | 1.65% | 0.26% |
|  | P7 | 17.14 Gb | 92.82% | 63.29% | 13.01% | 16.98% | 6.72% | 4.48% | 0.21% |
|  | P8 | 15.12 Gb | 94.11% | 64.79% | 11.40% | 18.74% | 5.07% | 2.86% | 0.20% |
|  | P9 | 22.05 Gb | 94.18% | 66.73% | 9.88% | 19.01% | 4.39% | 2.28% | 0.24% |
|  | P10 | 22.49 Gb | 91.43% | 63.18% | 11.34% | 17.96% | 7.53% | 5.59% | 0.24% |
|  | P11 | 20.00 Gb | 92.17% | 63.18% | 12.19% | 17.79% | 6.83% | 4.40% | 0.22% |
|  | U1 | 16.43 Gb | 95.26% | 68.61% | 8.42% | 20.06% | 2.92% | 1.29% | 0.12% |
|  | U2 | 14.34 Gb | 95.68% | 69.92% | 8.30% | 19.13% | 2.66% | 1.02% | 0.15% |
|  | U3 | 33.44 Gb | 95.39% | 69.00% | 8.11% | 20.20% | 2.68% | 1.08% | 0.14% |
| Ribo-m<br>inus | P1 | 15.46 Gb | 88.13% | 30.47% | 45.83% | 11.81% | 11.88% | 7.31% | 0.27% |
|  | P2 | 18.00 Gb | 90.09% | 25.86% | 53.97% | 9.33% | 10.8% | 5.83% | 0.25% |
|  | P3 | 19.71 Gb | 90.41% | 28.97% | 51.23% | 10.07% | 9.73% | 4.77% | 0.28% |
|  | P4 | 21.44 Gb | 93.75% | 25.55% | 57.57% | 9.42% | 7.46% | 2.16% | 0.28% |
|  | P5 | 17.41 Gb | 88.95% | 28.72% | 49.59% | 10.35% | 11.35% | 6.46% | 0.26% |
|  | P6 | 19.98 Gb | 92.86% | 28.62% | 54.76% | 9.55% | 7.07% | 1.83% | 0.34% |
|  | P7 | 17.12 Gb | 89.71% | 28.05% | 50.76% | 9.80% | 11.39% | 6.60% | 0.27% |
|  | P8 | 25.29 Gb | 91.85% | 31.76% | 50.32% | 10.31% | 7.61% | 2.65% | 0.33% |
|  | P9 | 24.95 Gb | 92.49% | 26.48% | 56.20% | 9.31% | 8.02% | 2.80% | 0.39% |
|  | P10 | 24.09 Gb | 93.01% | 23.67% | 59.62% | 8.39% | 8.32% | 2.83% | 0.40% |
|  | P11 | 29.93 Gb | 94.05% | 23.76% | 60.42% | 8.44% | 7.38% | 1.81% | 0.32% |
|  | U1 | 21.30 Gb | 94.61% | 37.72% | 41.81% | 13.90% | 6.58% | 0.96% | 0.23% |
|  | U2 | 18.24 Gb | 94.56% | 37.40% | 42.27% | 13.73% | 6.60% | 0.92% | 0.23% |
|  | U3 | 15.72 Gb | 94.19% | 36.81% | 42.57% | 13.77% | 6.86% | 1.19% | 0.26% |

### 2. Comparison of gene expression across protocols

To verify the reproducibility of the assay, we first evaluated the correlation of gene expression levels among all samples in two protocols by determining the ln (TPM + 1). We verified interbatch repeatability across 11 patient samples and intrabatch repeatability across 3 UHRR replicates.

Both the polyA+ and ribo-minus data showed high correlation in intrabatch repeatability, with r^2^values higher than 99%, as well as r^2^ values higher than 97% in interbatch repeatability (**Figure S1**). With good reproducibility, we compared the quantification of the 57,783 annotated genes in Gencode 43 by biotype in each method (**Figure 2A, Figure S2**). For 20,332 protein-coding genes, the total TPM in polyA+ data was 922,665, compared to 220,000 in ribo-minus data. This indicated that more reads could be used to calculate the expression of protein-coding genes and OMIM genes in the polyA+ method, which is consistent with the result that more unique reads were mapped onto exonic and exon–intron junction regions in the polyA+ method.

**Figure 2.**
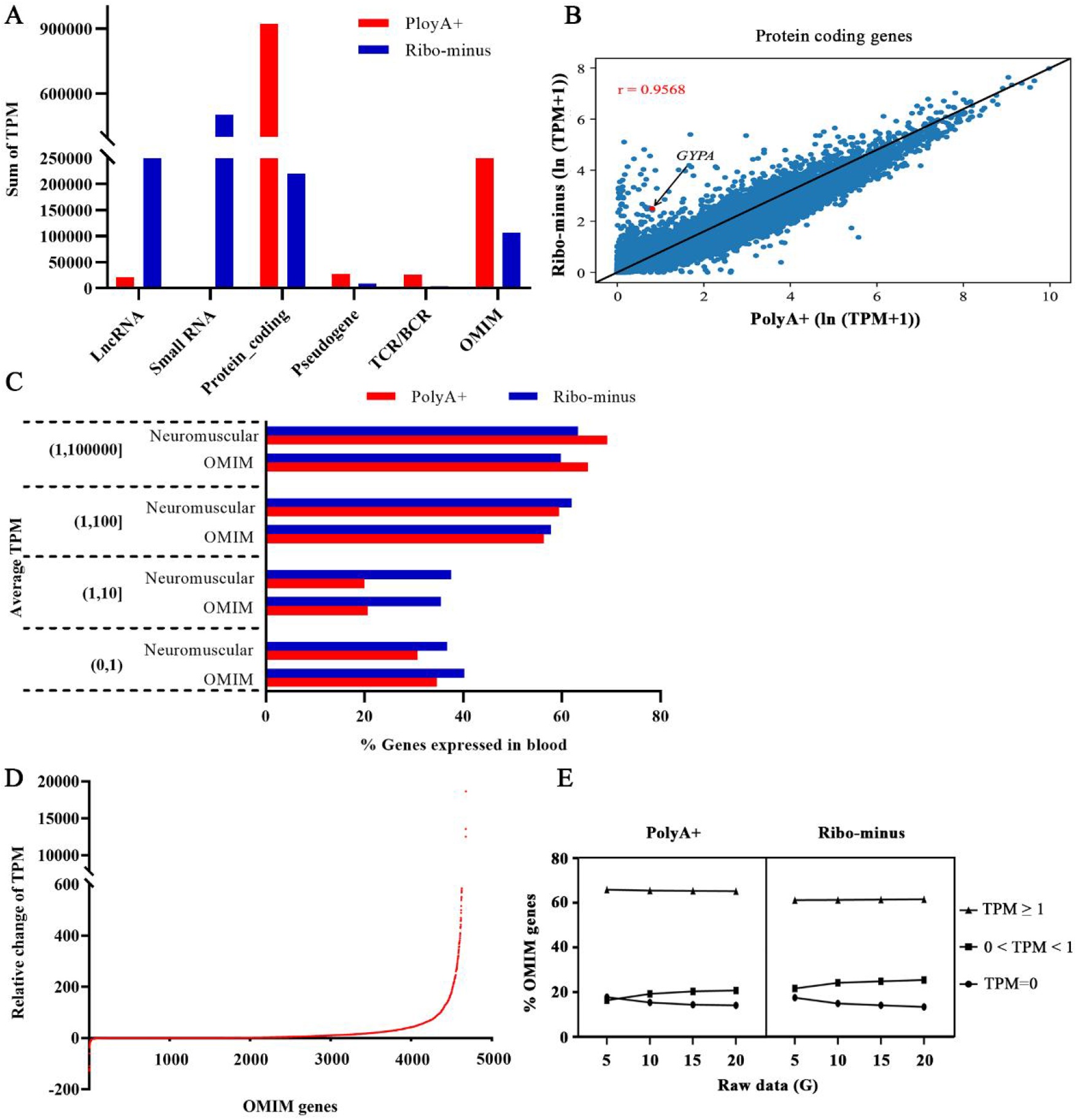
Comparison of gene expression. **(A)** Quantification of the 57,783 annotated genes in Gencode 43 by biotype. **(B)** The concordance of the protein-coding genes between the two methods. OMIM genes with exceptionally high expression (TPM ≥ 1) and large differences in results between the two protocols (log2FC (fold change) > 4) are labelled in red. **(C)** The proportion of OMIM genes in different TPM ranges. **(D)** Relative changes in TPM values of 4,678 OMIM genes. Compared to that in the rRNA depletion (ribo-minus) data, the median TPM of OMIM genes in polyA-selection (polyA+) data increased by 3.435. **(E)** OMIM gene expression among different sequencing bases (G, Gb). The percentage of OMIM genes with TPM > 1 changed little among four groups.

Although the overall concordance was high, it clearly varied from biotype to biotype. We next compared the concordance of gene expression between the two methods by determining the ln (TPM + 1) of a single gene. Among different biotypes, the correlation coefficient of the protein-coding genes between the two methods was 0.9568 in the blood (**Figure 2B, Figure S3**) albeit that the polyA+ method had a bias in sequence coverage across genes, with poor coverage at the 5′-end, especially for genes with longer transcripts (**Figure S4**). Very few genes were detected uniquely in the polyA+ data, whereas many more genes were detected only in the ribo-minus data. This is probably because the ribo-minus protocol should capture both polyA+ and polyA-transcripts, whereas the polyA+ selection protocol should almost exclusively detect transcripts with polyA+ tails. Among genes with exceptionally high expression (TPM ≥ 1) and large differences between the two protocols (log2FC (fold change) > 4), we evaluated whether they were OMIM genes and labelled them red if they were in OMIM (**Figure 2B**).

The expression levels of OMIM genes also needs attention and comparison since genes with low or no expression in blood cannot be used for clinical detection. Therefore, we calculated the proportion of genes in different TPM ranges. Overall, 65.29% of OMIM genes were expressed in blood (TPM > 1(12)) in polyA+ data, while only 59.79% were expressed in the ribo-minus data. Specifically, the majority of genes had TPM values in the range of 1-100 in the polyA+ data, whereas most genes had TPM values in the range of 1–10 or below 1 in the ribo-minus data (**Figure 2C**). Based on the average TPM value of the 4,678 OMIM genes in ribo-minus data, we calculated the relative changes in TPM corresponding to the polyA+ data (**Figure 2D**). It is clear that the TPM value of most genes increased, and the median TPM value increased by 3.435. This means that a median of 258 more valid reads could be analysed per gene. Next, to investigate whether increasing sequencing bases can improve OMIM gene expression, we selected polyA+ data and ribo-minus data of P9 sample and extracted different sequencing bases into four groups. In **Figure 2E**, the percentage of OMIM genes with TPM > 1 changed little among four groups. The proportion of OMIM genes with TPM between 0 and 1 increased along with the proportion of OMIM genes with TPM =0 decreased. Thus, genes with low expression (TPM < 1) are more sensitive to sequencing bases, and the improvement of gene expression is limited with the increased sequencing bases. Therefore, the polyA+ method with at least 5 Gb sequencing data is recommended for clinical detection, and genes with TPM ≥ 1 are recommended to be listed as detectable genes.

In general, although both the polyA+ and ribo-minus data showed high reproducibility, and although there was a high correlation of the protein-coding gene expression between the two methods, the expression level of the OMIM genes was higher in the polyA+ data, with a median of 258 more valid reads that could be used for analysis per gene. In addition, the proportion of OMIM genes with TPM > 1 increased by ∼5%, which means that more OMIM genes can be accurately quantified in polyA+ data, leading to a larger range of detectable genes.

### 3. Differentially expressed genes

Control data have a great impact on differential gene expression analysis, and different pipelines have different requirements for input data. For example, EdgeR requires that the patient group must have the same number of samples as the control group, whereas DROP analysis does not have this requirement. When RNA-seq is used as a clinical test, it is more typical and also convenient to compare an individual test sample with other controls. Therefore, we took the average gene expression of the remaining sample data in the corresponding protocol as the control, used EdgeR to analyse the data for samples individually, took the remaining sample data from the corresponding protocol as a whole control, and used DROP to analyse the samples individually. Then, we determined the concordance of differentially expressed genes in polyA+ data and ribo-minus data and evaluated the burden for subsequent interpretation. For genetic diagnosis, we paid more attention to differentially expressed OMIM genes. An OMIM gene was defined as differentially expressed if (1) its fold change > 2, (2) its p value < 0.05, and (3) the mean expression ≥ 1 TPM. The total number of differentially expressed OMIM genes analysed by EdgeR and DROP in each protocol is shown in **Figure 3A**. In our results, the median number of differentially expressed OMIM genes analysed by DROP was less than that analysed by EdgeR regardless of which experiment was performed, and the number varied greatly across samples when using EdgeR. There was no significant difference in the number of downregulated DEGs between polyA+ and ribo-minus regardless of whether EdgeR or DROP was used, whereas the number of total DEGs and the upregulated DEGs between polyA+ and ribo-minus was significant when using EdgeR (p = 0.032 and p = 0.0098, paired samples Wilcoxon signed rank test). In addition, there was no significant difference in the number of DEGs in each method when analysed by either EdgeR or DROP except for the upregulated DEGs in polyA+ data when analysed by EdgeR. These results indicated that the number of DEGs varies greatly between methods because the control set of EdgeR is averaged as one sample, resulting in the difference in the numbers of DEGs. To further compare the DEGs of different combinations, the overlap of the DEGs identified per sample in four combinations was shown, classified as upregulated **(Figure S5**) and downregulated (**Figure 3B**); downregulated DEGs are more common in genetic disease diagnosis. We believe that genes detected as aberrantly expressed in all four combinations are highly likely to be true positives, while the genes detected in only one combination may be false positives attributable to method specificity. In our results, we can see that few differentially expressed genes were detected in all four combinations. Only 1 in P5 and 2 in P11 were detected, and most of them were detected unique to different combinations. In this case, higher expression levels were more conducive to reducing false positives, and DROP was more stable than EdgeR. Therefore, the combination of polyA+ and DROP would yield higher specificity.

**Figure 3.**
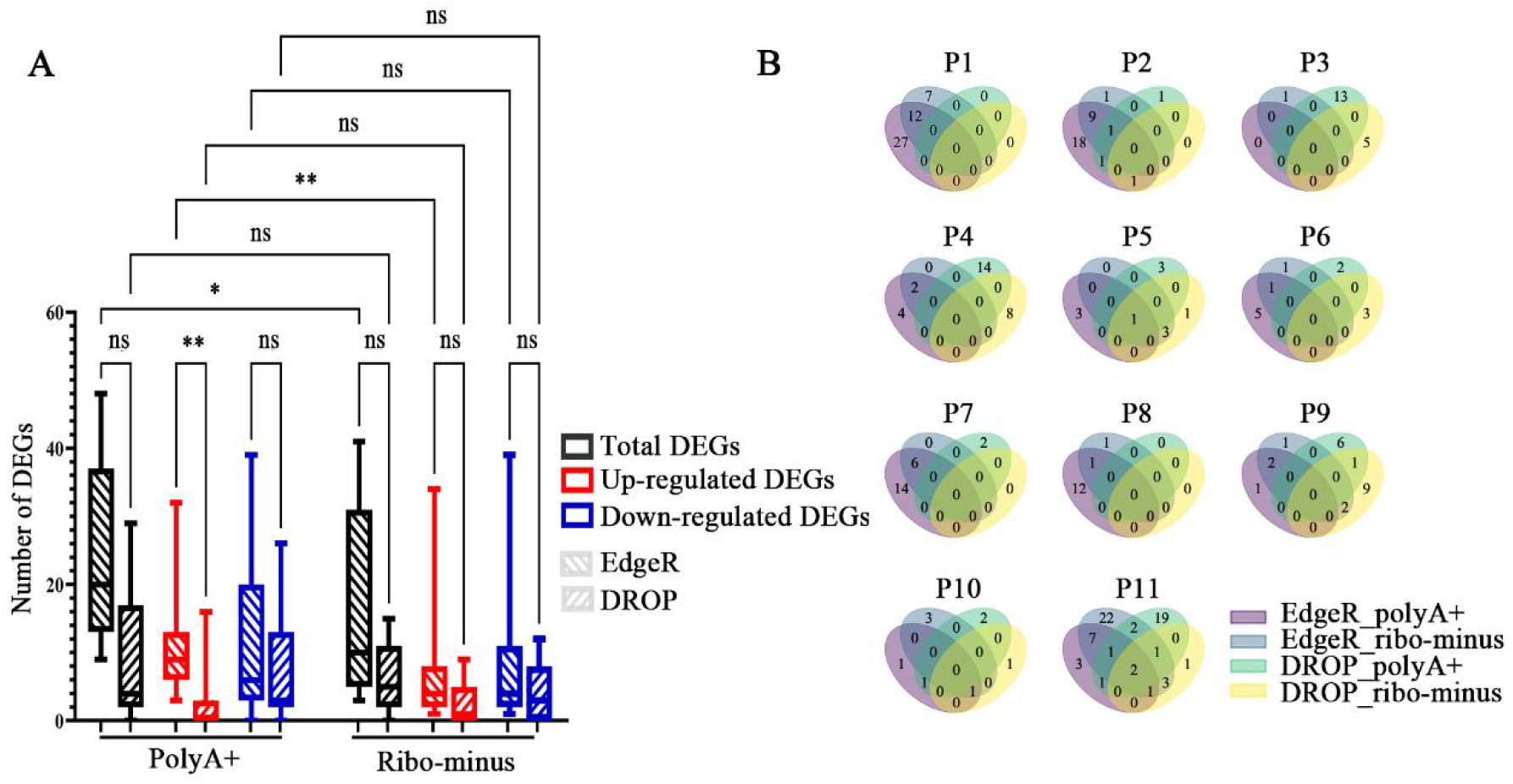
Comparison of differential gene expression. **(A)** Box plot of the number of differentially expressed genes (DEGs) identified by EdgeR and DROP in polyA+ data and ribo-minus data from 11 patients. The number of subgroups including upregulated DEGs and downregulated DEGs in different combinations is also shown. Paired samples Wilcoxon signed rank test was used for between-group analyses, ^*^ p < 0.05, ^**^ p < 0.01, ns p > 0.05. **(B)** The overlap of downregulated DEGs identified with different combinations of experimental methods and bioinformatic pipelines in each patient.

### 4. Accuracy in splicing changes

Leafcutter and rMATS were simultaneously used for analysing the alternative splicing events in polyA+ data and in ribo-minus data with the remaining sample data from the corresponding protocol as the control dataset, DROP was not used because it requires at least 30 samples as control dataset. Aberrant splicing was defined if (1) its p value was < 0.05; (2) the mean expression TPM ≥ 1, and (3) the gene was an OMIM gene. In **Figure S6**, the median number of aberrant splicing events and genes analysed by Leafcutter is less than that analysed by rMATS regardless of which experiment was performed. One reason is that Leafcutter has limitations in detecting intron retention events. There was a significant difference between polyA+ data and ribo-minus data in terms of the number of aberrant splicing events identified by using either Leafcutter or rMATS (p = 0.001 and p = 0.002, paired samples Wilcoxon signed rank test). Similarly, the median number of aberrant splicing events or genes in ribo-minus data was less than that in polyA+ data regardless of which software was used for analysis.

To compare the accuracy of the two methods in detecting splicing changes, we combined the RNA-seq and genomic data and screened those variants occurring in or near the aberrant splicing region with a spliceAI prediction score greater than 0.5 and minor allele frequency ≤ 1% for verification. Finally, 3 variants involving aberrant splicing events were selected for RT–PCR validation. Three variants and their primer sequences are shown in **Table S1**. For each group, one or two samples from individuals of the same sex and a similar age were used as controls. The sashimi plots of the polyA+ data and ribo-minus data, with corresponding RT-PCR results, are shown in **Figure 4**. In polyA+ data, an aberrant splicing event in the *COPB1* was missed by Leafcutter and rMATS, while it was identified by Leafcutter in ribo-minus data with only one read supporting this alternative splicing. In fact, the threshold of reads supporting splicing is ≥ 5 (11) or ≥ 20 (12) according to previous studies, and the 3 variants would be missed when using the ribo-minus data due to filtering. Although the number of verified splicing variants in this study is limited, it can be inferred that the ribo-minus data may yield more false positives despite detecting more aberrant splicing events without junction read filtering. In contrast, when poly A+ data was used, junction reads supporting the other two variants were significantly more numerous, and one variant was missed. Therefore, in the case of aberrant splicing, different performances were presented by the two methods depending on the read filtering conditions applied. The combination of polyA+ with read filtering is recommended for clinical use in splicing detection due to its higher sensitivity and specificity.

**Figure 4.**
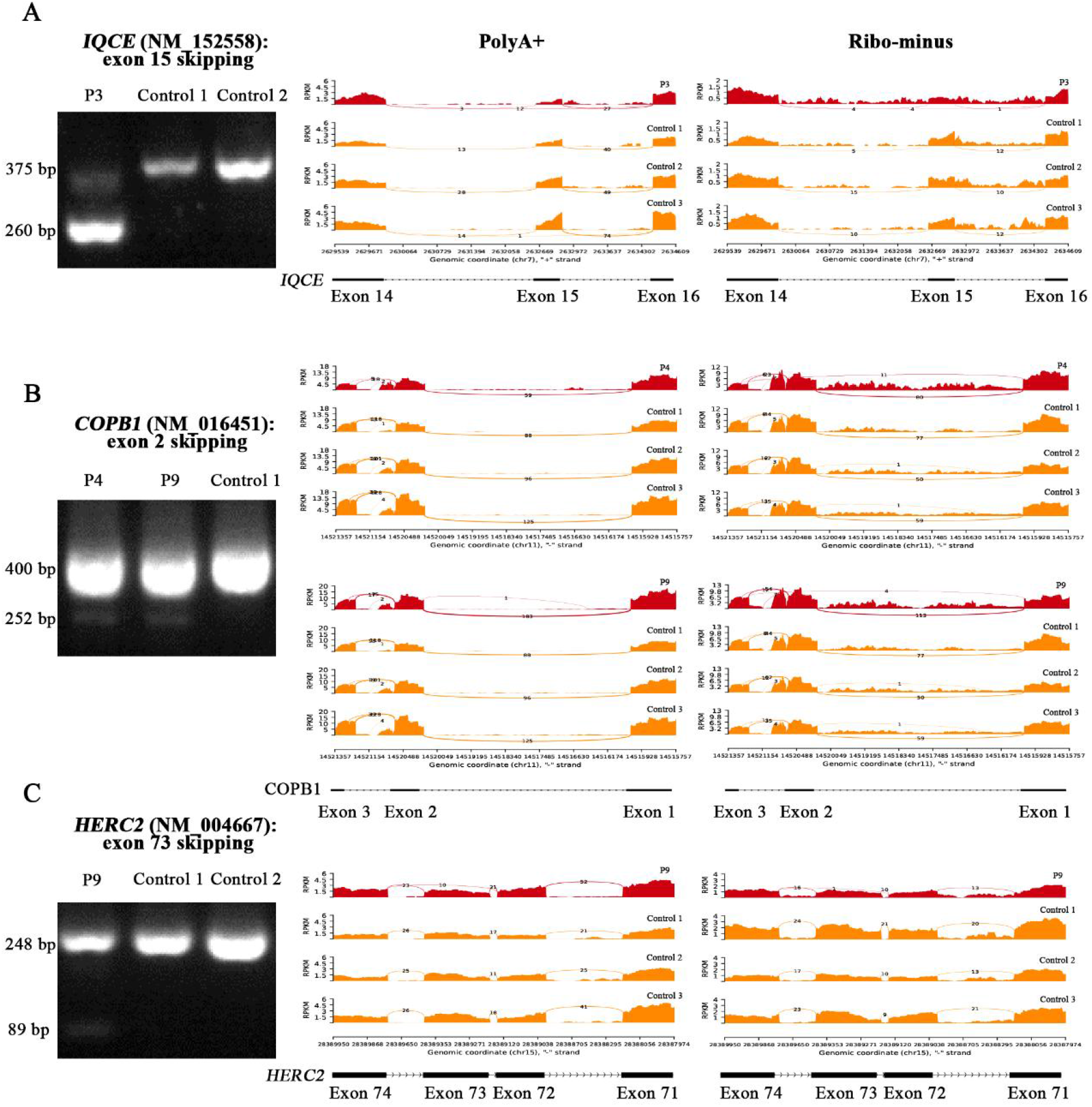
Comparison of aberrant splicing. Sashimi plot of the polyA+ data and ribo-minus data with corresponding RT-PCR results for **(A)** *IQCE* (NM_ 152558.3): c.1349+1G>A; **(B)** *COPB1* (NM_ 016451.4): c.-57-2A>G; **(C)** *HERC2* (NM_ 004667.5): c.11141-2del.

### 5. Monoallelic expression of rare variants

RNA-seq data can be used to identify expressed rare variants in which only 1 of the 2 copies of a gene is active while the other is silent. DROP was used for MAE analysis, and a variant was defined as MAE if (1) the variant was covered by more than 10 reads; (2) the Hochberg adjusted p value < was 0.05; (3) the altered allele frequency was ≥ 0.8; (3) the minor allele frequency was < 0.001; and (4) the molecular basis of the variant-associated genes in the OMIM database was definite. Whole-genome sequencing data from ten patients were used for MAE analysis. In the polyA+ data and ribo-minus data, the median number of MAE variants identified was 14.5 and 1.5, respectively (**Figure 5A, Figure 5B**). To further compare the MAE variants in these two methods, the overlap of the MAE variants identified per sample in polyA+ data and ribo-minus data is shown in **Figure 5C**. Six out of ten samples had overlapping MAE variants, and all of these MAE variants were in intronic or untranslated regions, except for one in the exon region. However, no further interpretation was performed because diagnosis was not our aim in this case.

**Figure 5.**
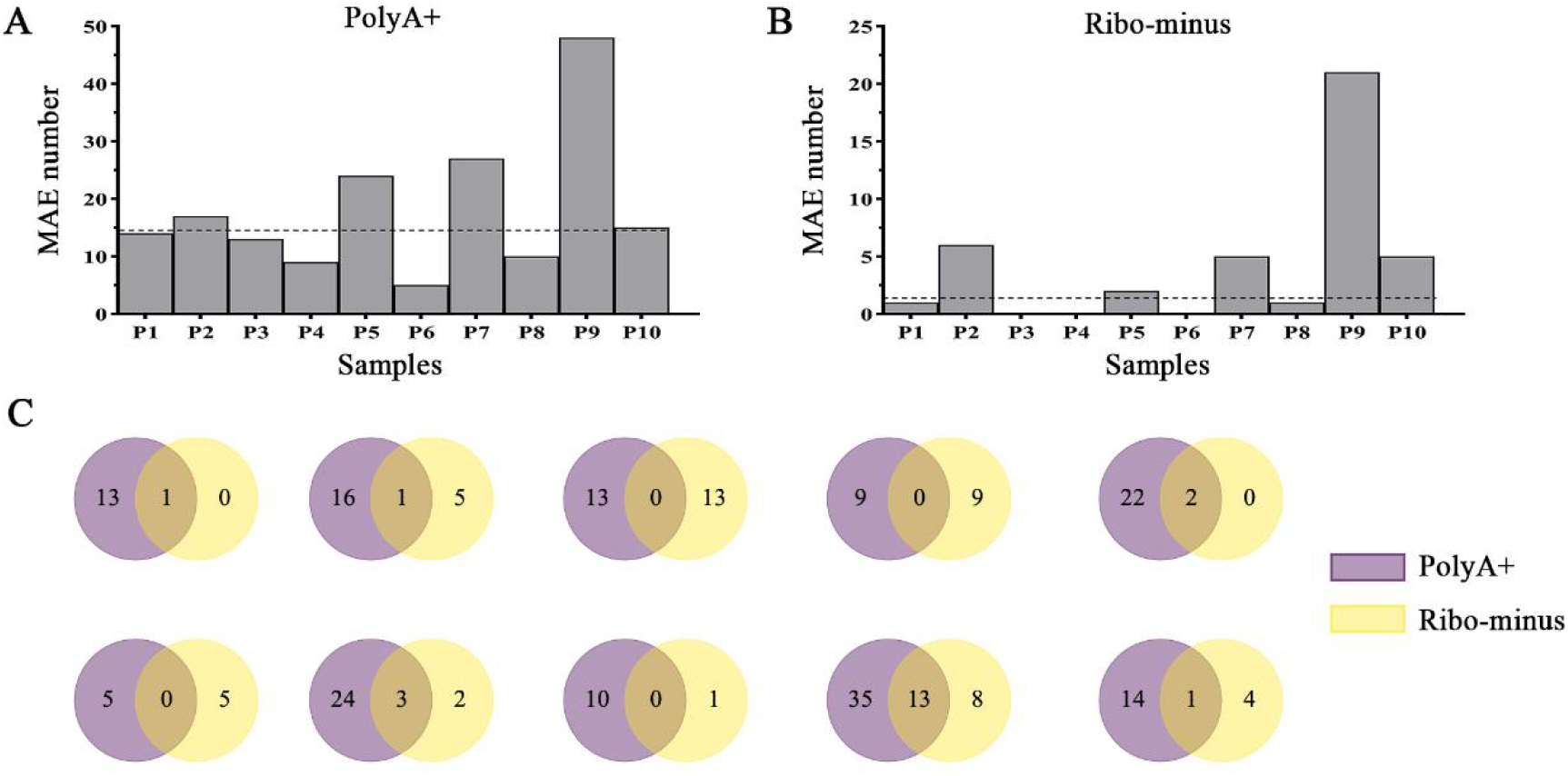
Comparison of monoallelic expression of rare variants. **(A)** Bar chart of the identified monoallelic expression (MAE) of rare variants across samples in the polyA+ data. The median number of identified MAE variants is 14.5. **(B)** Bar chart of the identified MAE of rare variants across samples in the ribo-minus data. The median number of identified MAE variants is 1.5. **(C)** The overlap of MAE variants identified using the two experimental methods in each patient.

## Discussion

Altogether, our results confirm that the polyA+ method is more conducive to Mendelian disease diagnosis when using blood-based RNA-seq in clinical practice. We demonstrate that a higher proportion of unique reads from the polyA+ library were mapped to exonic and exon– intron junction regions (84.54% vs. 40.14%), resulting in more detectable OMIM genes with TPM > 1 in the blood (65.29% vs. 59.79%). The mapping of more reads to intronic regions in ribo-minus data means that a median of 258 more valid reads per gene are needed to achieve the same level of accurate exon/gene quantification as is provided by polyA+ data, necessitating higher sequencing costs. Moreover, although the transcriptome profiling of protein-coding genes in the two methods are highly correlated, polyA+ is more sensitive to aberrant splicing under common filtering conditions, which is important for evaluating variants of uncertain significance for pathogenicity, and more sensitive to MAE variants, which elevates the diagnostic yield. Finally, the combination of polyA+ and DROP was recommended to obtain higher sensitivity and specificity. Overall, the combination of polyA+ and DROP is recommended when implementing blood-based RNA-seq for the diagnosis of Mendelian diseases in clinical practice, and filtering criteria for DEGs, AS and are suggested for reference.

When implementing RNA-seq as an adjunct to DNA sequencing in rare disease clinical diagnostics, many factors should be considered in addition to analytical validity, including clinical validity, clinical utility, cost and turnaround time (TAT)(25). For example, we noted that the operation of the modified polyA+ method is simpler than that of the ribo-minus method, with a shorter processing time per sample by an average of 0.5 hours. This may be insignificant in the overall TAT. However, the effect is noticeable when the sample size is large, and the polyA+ approach is easier to automate. In addition, polyA+ has lower reagent costs than ribo-minus, as well as lower sequencing costs to achieve the same exonic coverage. However, regardless of which method is used, the clinical utility of RNA-seq for rare disease clinical diagnostics is beyond doubt. More data are needed to clarify whether there is a difference in the clinical validity of these two methods.

Although previous studies have consistently recommended the polyA+ method for eukaryotic mRNA studies, molecular diagnostic RNA-seq has largely been conducted in the research arena. Systematic evaluation of this technology is needed before it can be used in the clinic. Our study has provided data to allow the comparison of analytical validity between the two commonly used blood RNA-seq approaches, with a few limitations. On the one hand, the number of samples used for comparison was small, and the patient samples did not carry known MAE variants or differentially expressed genes to allow validation of the method accuracy. Moreover, similar to the utility of large databases of control exomes for Mendelian disease diagnosis (26, 27), a large control transcriptome dataset derived from the same tissue, ideally using the same RNA selection and library preparation protocols and sequenced on the same platform, is required for RNA-seq analysis (12, 13, 15). However, in our study, we used the remaining 10 sample data from the corresponding protocol as controls without introducing an external dataset such as GTEx due to the following considerations. First, blood transcriptome data in GTEx were produced using a standard non-strand-specific protocol with polyA selection of mRNA, which is different from our RNA selection and library preparation methods, especially the ribo-minus method. Therefore, inaccurate evaluation and comparison could result from biases inherent in different methods.

Secondly, the causal defects differ between patients, which is reasonable for Mendelian diseases with a diversity of 4000+ known disease-causing genes, so the patients serve as good controls for each other. Since the control dataset has a significant impact on the accuracy of the test, a minimum of 30 samples is needed for empirical detection (28), and a pipeline to generate matching controls over time should be included during clinical application.

Implementation of RNA-seq in clinical diagnostics requires establishment and standardization of methods for assessing analytical validity. Through blood RNA-seq, our study assessed two main methods of RNA selection for their different data performance in aberrant expression, aberrant splicing and MAE variants and provided some thresholds for filtering and interpretation criteria for reference. However, since gene expression is tissue specific, the combination of multiple “omics” sources is necessary for the further diagnosis of unsolved rare disease cases in the future regardless of which method is chosen.

## Data Availability

All data produced in the present study are available upon reasonable request to the authors

https://db.cngb.org/

## Declarations

### Ethics statement

Written informed consents were obtained from all the participants. This study was approved by the Ethics Committee of Peking Union Medical College Hospital (NO. JS-3130) and was performed in accordance with the Declaration of Helsinki.

## Authors’ contributions

J.S. and Y.D. designed the research. Z.Y. wrote the first draft of the article. X.Y. designed and performed the experiments. Y.C. and Z.W. performed data analysis, XZ.Y. recruited patients and collected specimens. L.S. and Z.P. contributed to drafting and revising the manuscript. All authors reviewed the manuscript and approved the submitted version.

## Data Availability Statement

The data that support the findings of this study have been deposited into CNGB Sequence Archive (CNSA)(29) of China National GeneBank DataBase (CNGBdb)(30) with accession number CNP0004191. The datasets for this article are not publicly available due to privacy or ethical restrictions. Requests to access the datasets should be directed to the corresponding author.

## Acknowledgments

We thank all the blood donors for their invaluable contribution to this study.

## Figures

**Figure S1.**
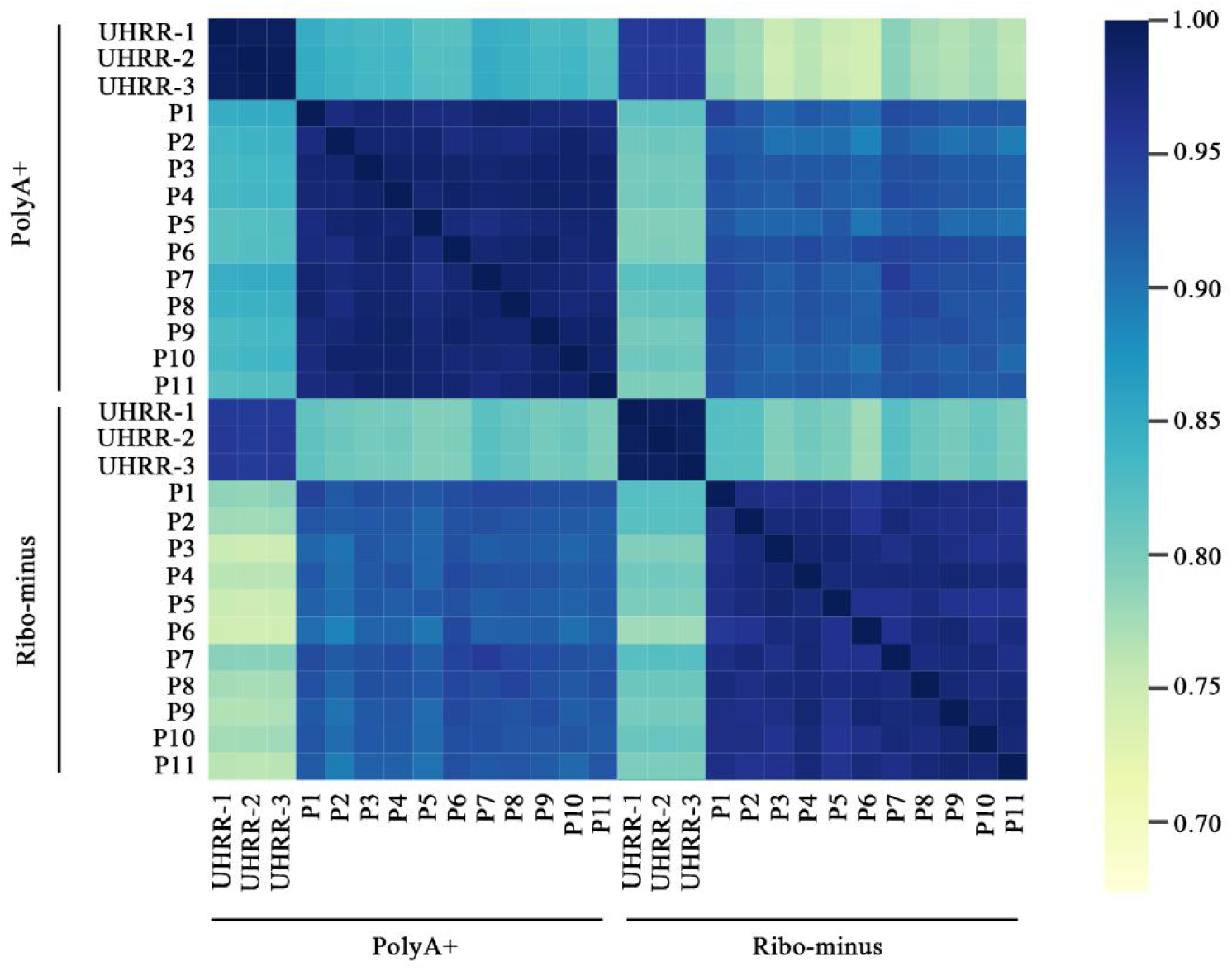
Correlation of gene expression levels in two protocols. Interbatch repeatability with 11 patient samples. The r^2^ was higher than 99% for both the polyA-selection (polyA+) data and rRNA depletion (ribo-minus) data. Intrabatch repeatability with 3 UHRR replicates. The r^2^ was higher than 97% for both the polyA+ data and ribo-minus data.

**Figure S2.**
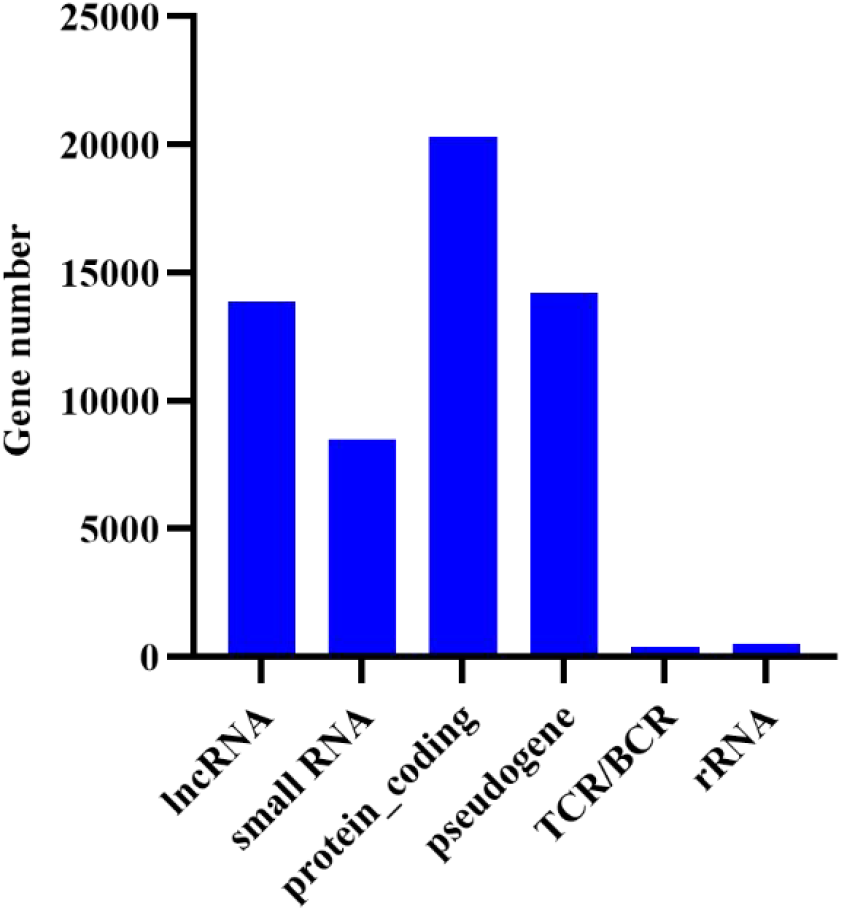
The biotypes of 57,783 annotated genes in Gencode 43. The largest number is protein-coding genes, with a total of 20,332.

**Figure S3.**
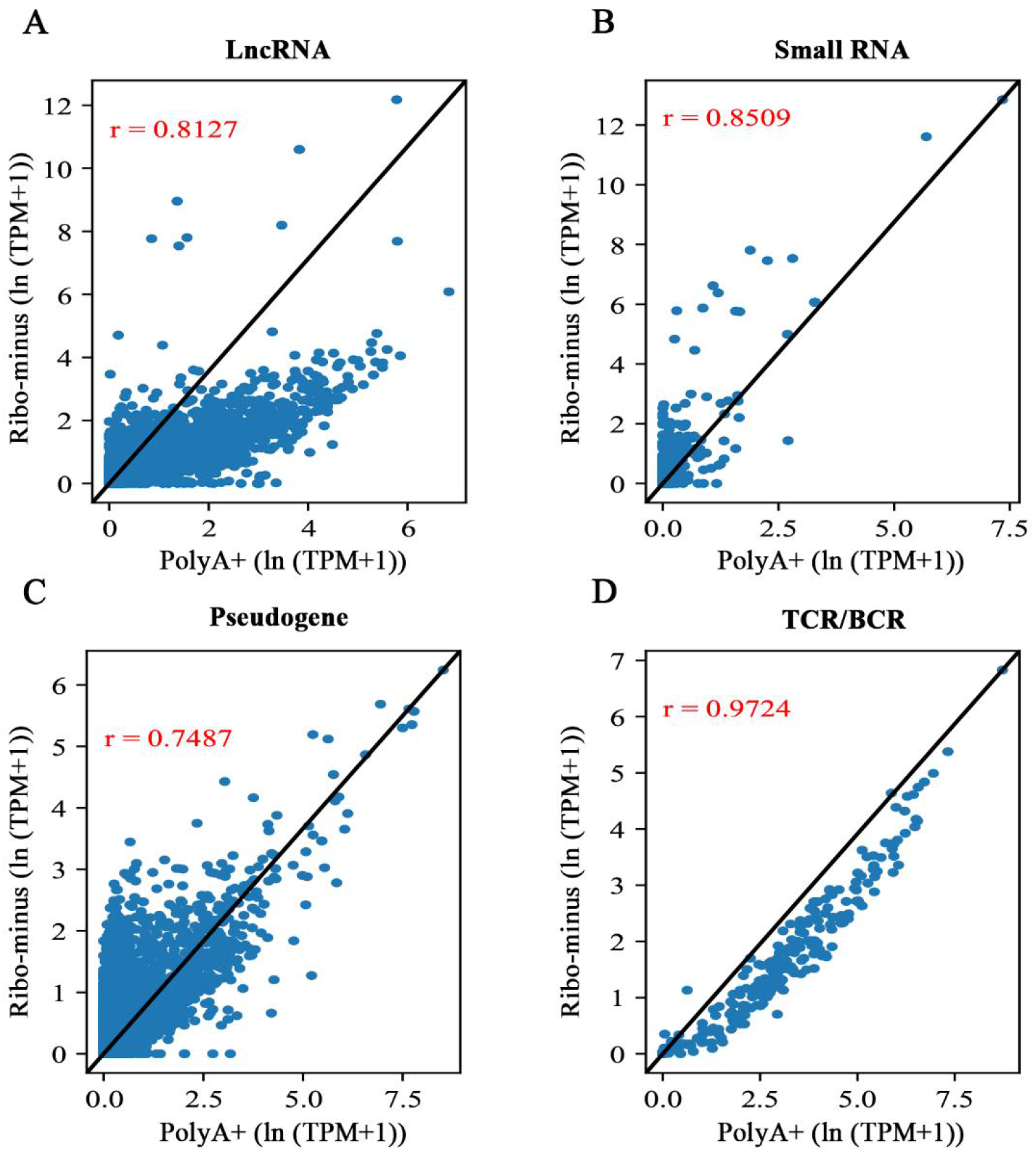
Concordance of gene expression between the polyA-selection (polyA+) and rRNA depletion (ribo-minus) methods by biotype. **(A)(B)(C)** and **(D)** represent lncRNA, small RNA, pseudogenes, B-cell receptors and T-cell receptors (TCR/BCR), respectively. The x- and y-axes indicate ln (TPM + 1). TPM is transcripts per million.

**Figure S4.**
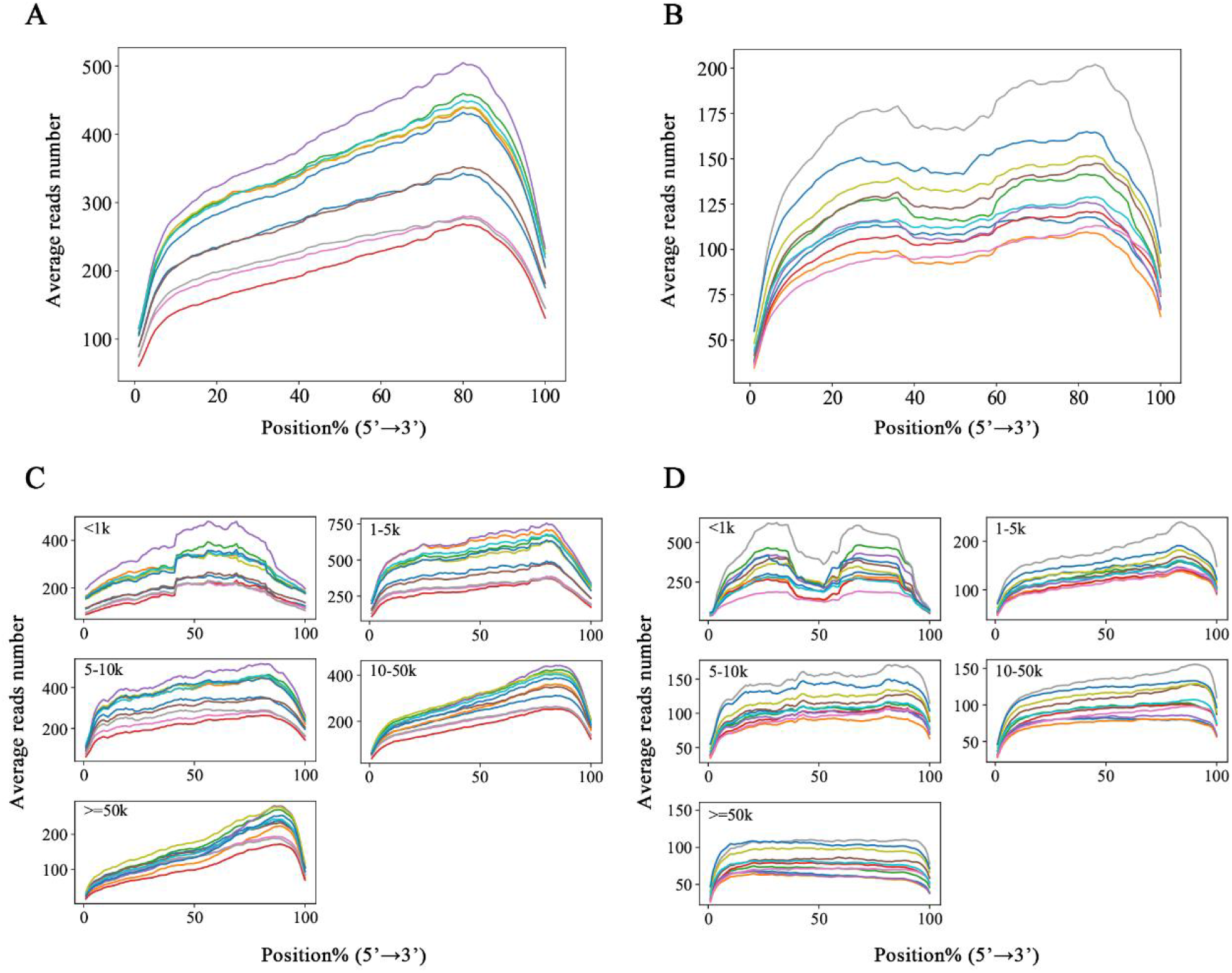
Read distribution across the gene body. Relative coverage in the **(A)** polyA-selection (polyA+) and **(B)** rRNA depletion (ribo-minus) methods. Samples with different coverage are indicated by colour. The coverage of the ribo-minus method is more uniform. We also plotted the length-dependent coverage for the **(C)** polyA+ and **(D)** ribo-minus data. Note that larger transcripts show a stronger distribution bias in the polyA+ dataset.

**Figure S5.**
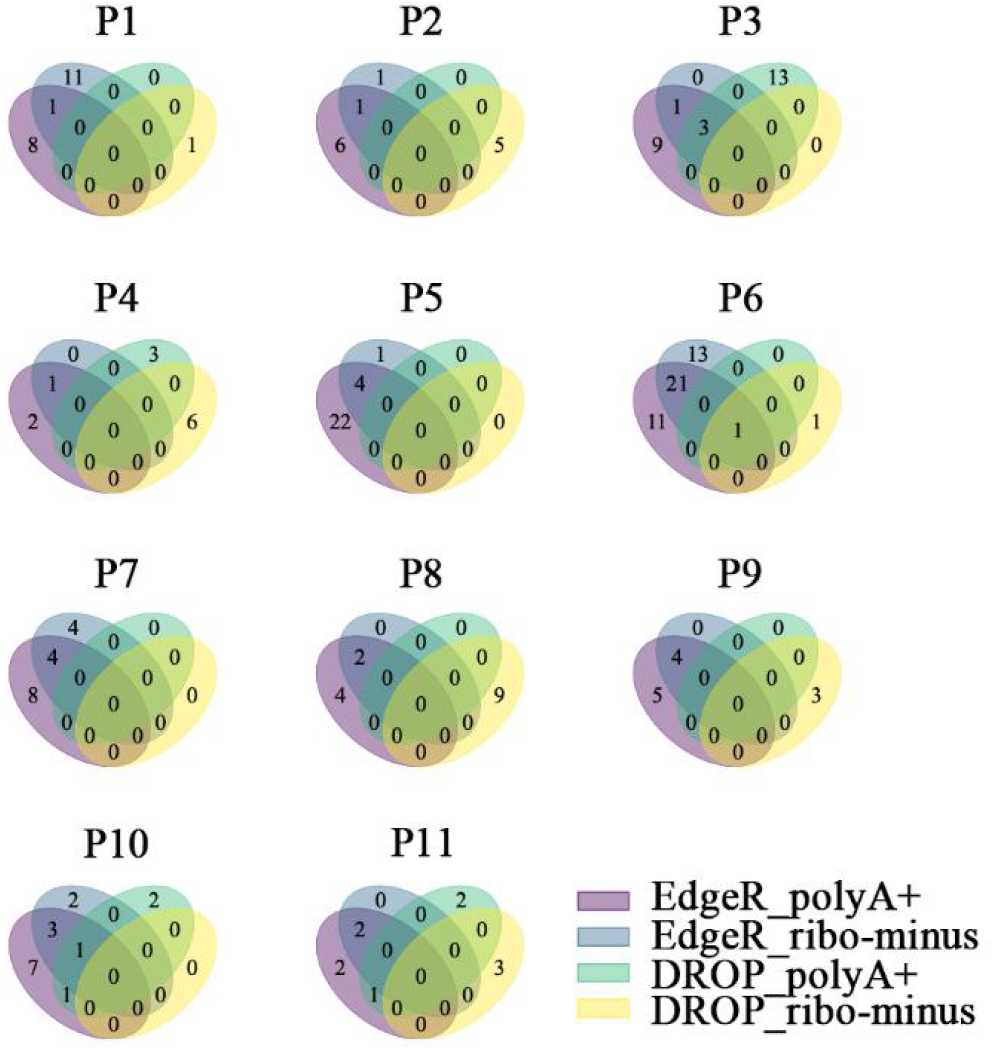
The overlap of upregulated DEGs in different combinations of experimental methods and bioinformatic pipelines in each patient.

**Figure S6.**
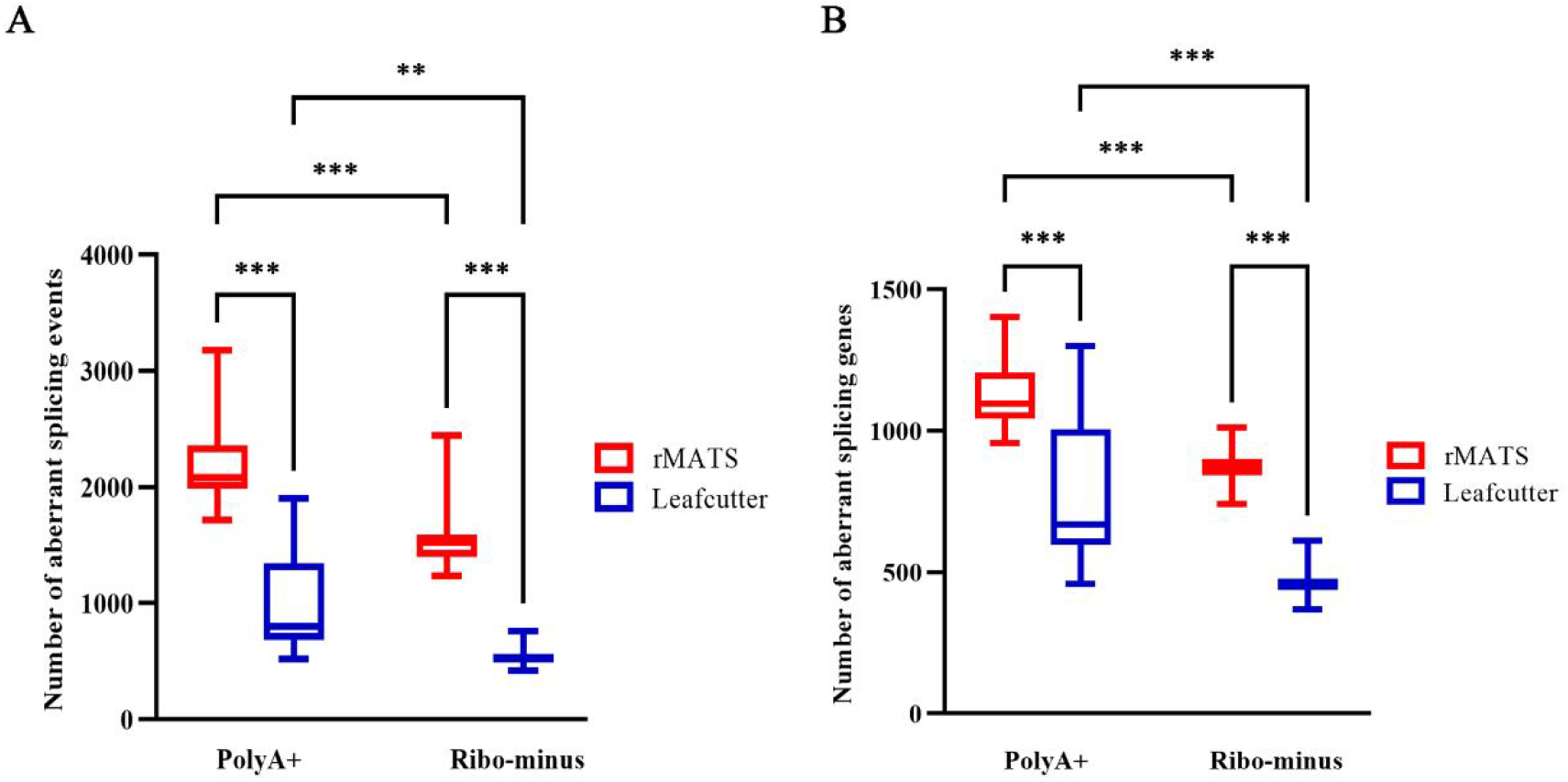
Comparison of aberrant splicing. Box plot of the number of **(A)** aberrant splicing events and **(B)** genes analysed by Leafcutter and rMATS in polyA-selection (polyA+) data and rRNA depletion (ribo-minus) data from 11 patients. The paired samples Wilcoxon signed rank test was used for between-group analyses, ^*^ p < 0.05, ^**^ p < 0.01, ns p > 0.05.

