## Supplementary figures and images for "Systematic evaluation of the two main blood-based RNA-seq approaches for Mendelian disease diagnosis"

### Supplemental Figure S1

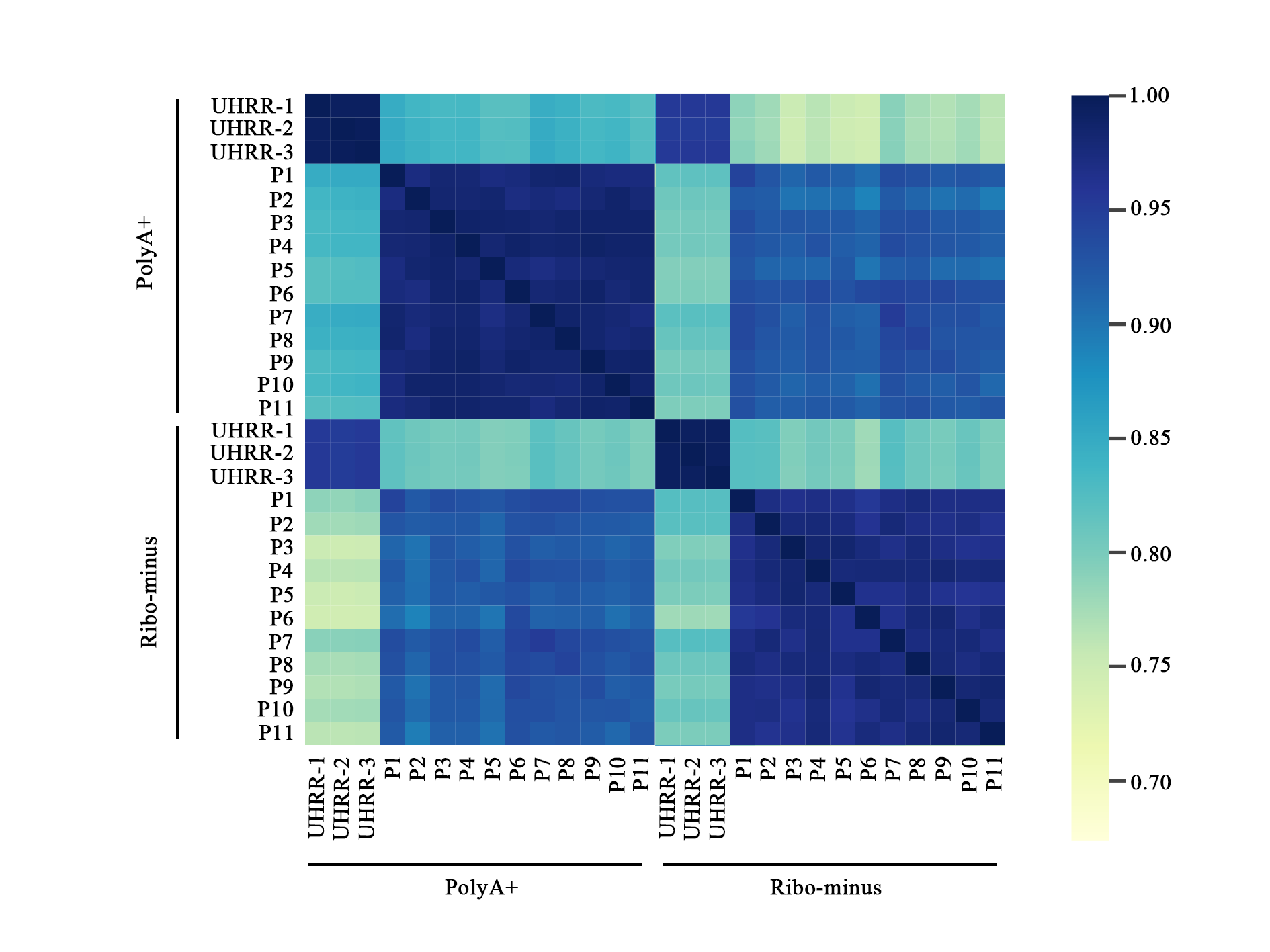

### Supplemental Figure S2

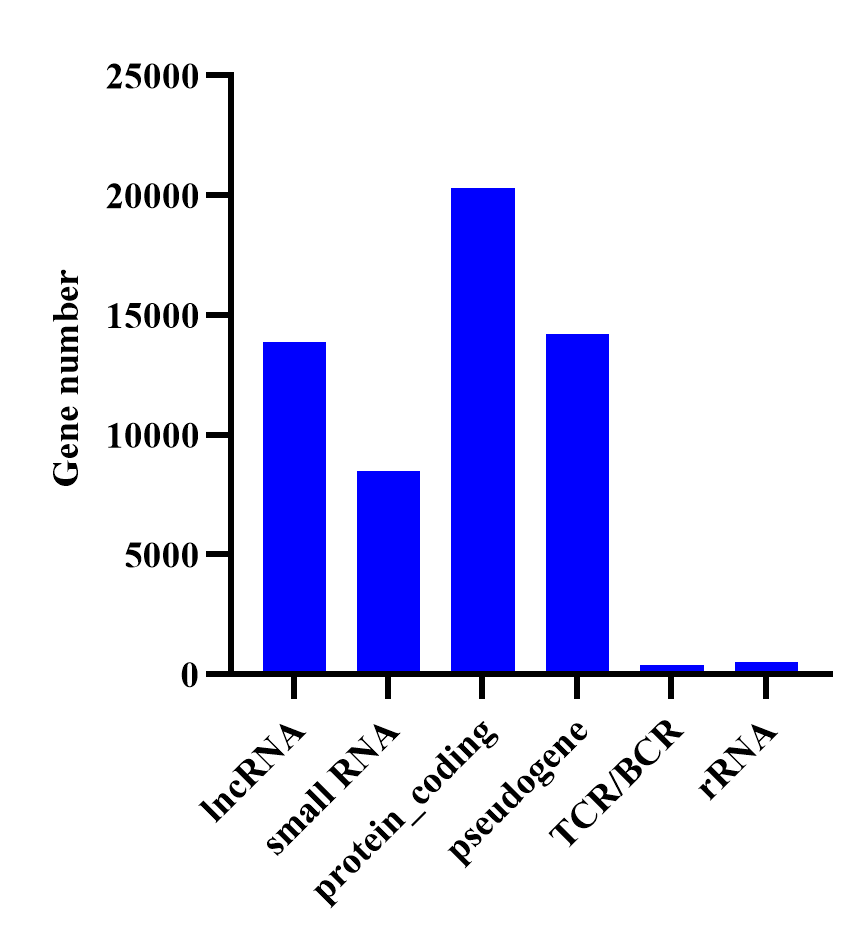

### Supplemental Figure S3

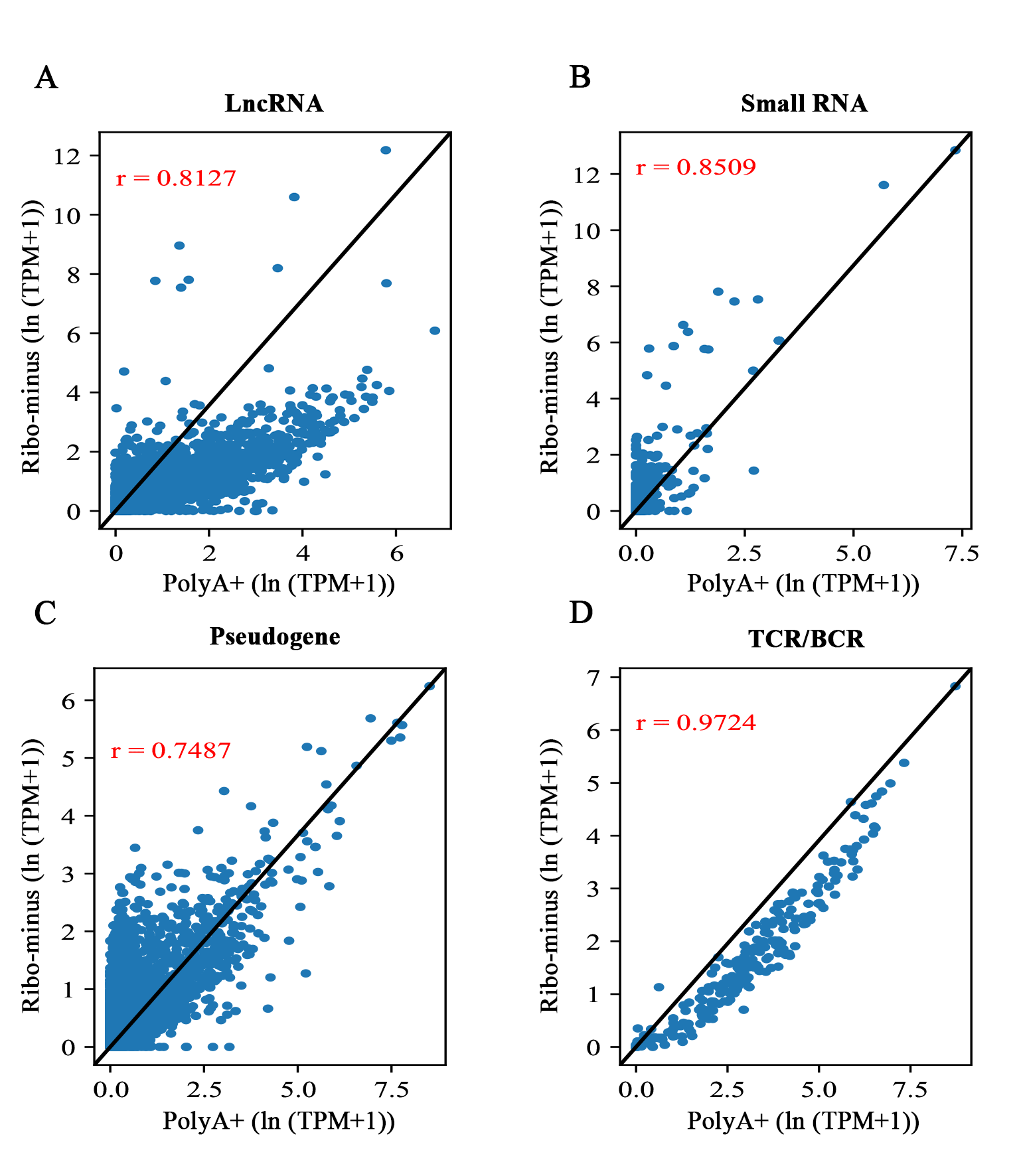

### Supplemental Figure S4

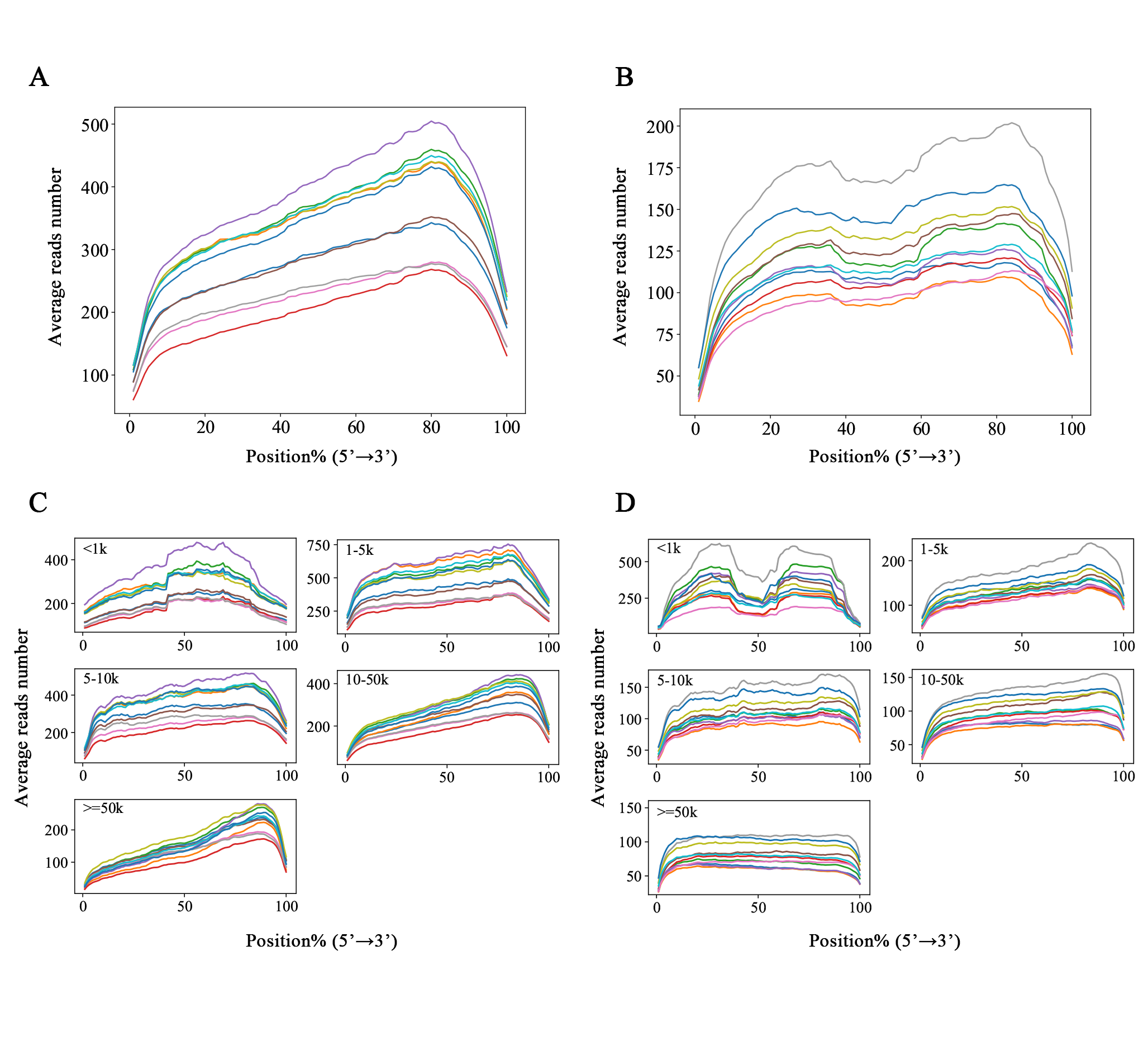

### Supplemental Figure S5

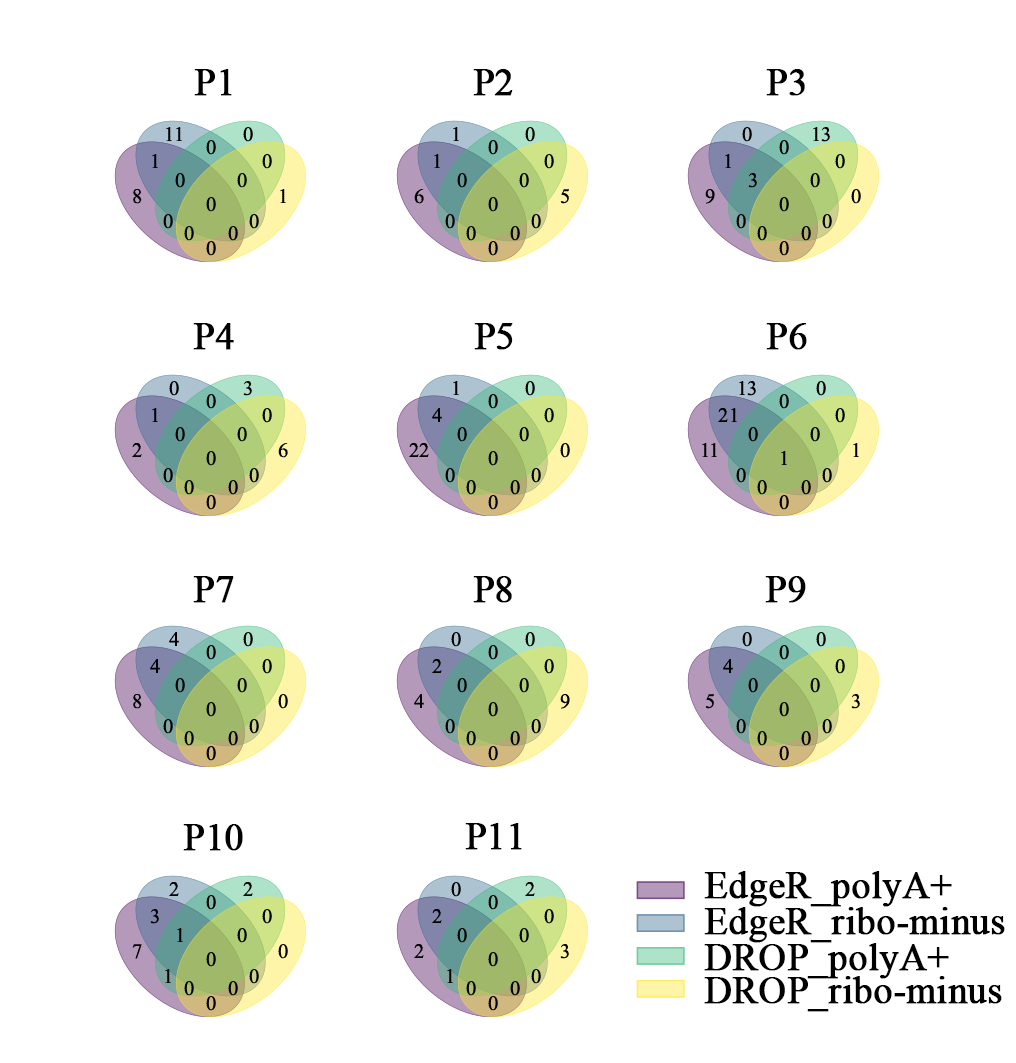

### Supplemental Figure S6

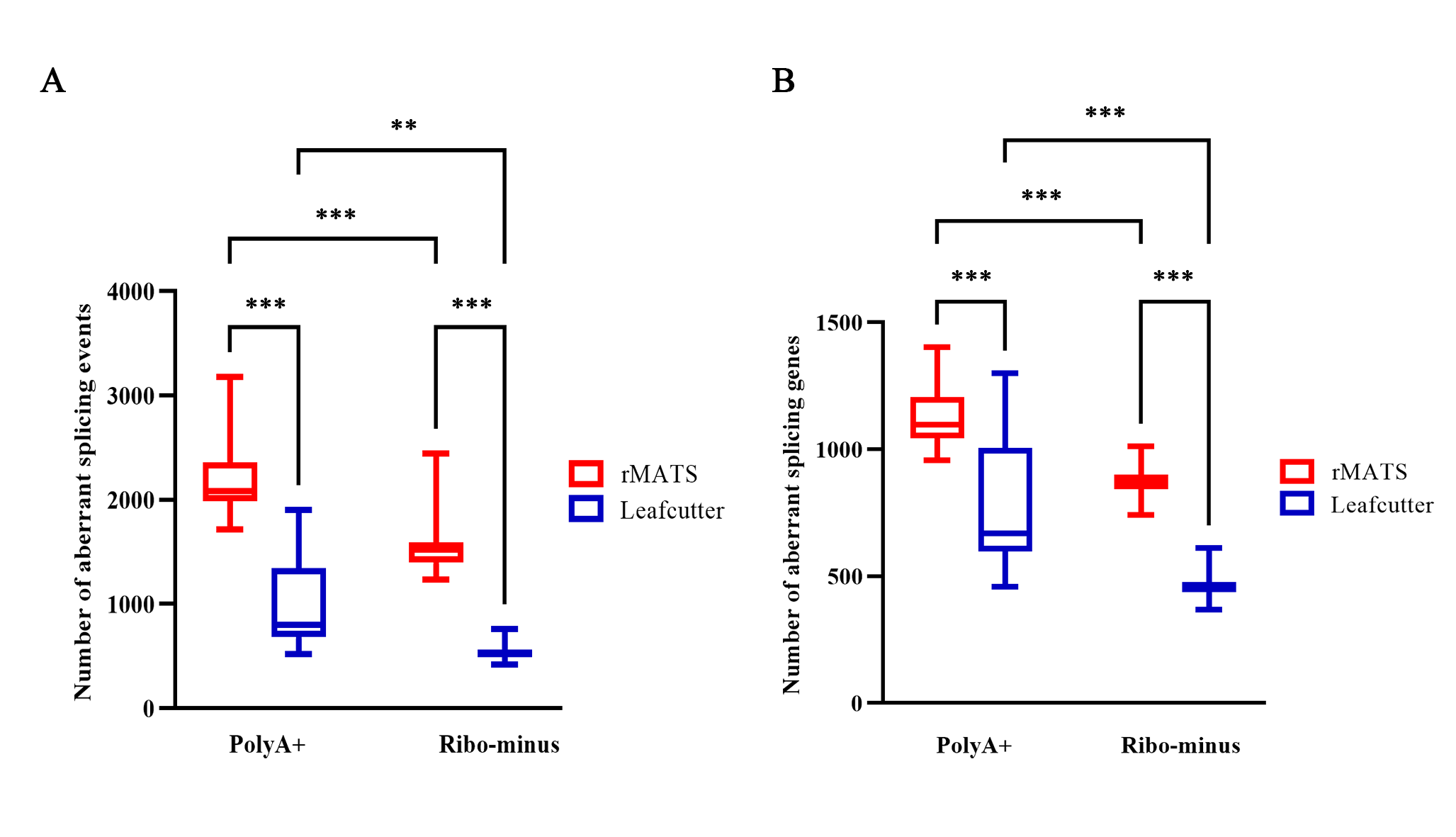
